# Comparison of mechanical homogenization versus enzymatic digestion sample preparation methodologies for SARS-CoV-2 detection in saliva for surveillance of variants of concern on the University of Tennessee campus in early 2021

**DOI:** 10.1101/2022.05.11.22274949

**Authors:** Magen R. Poindexter, Tingting Xu, Cynthia M. Swift, Caleb M Proctor, Fadime Kara-Murdoch, Zachary P Morehouse, Gabriella L Ryan, Frank E. Löffler, Rodney J Nash

**Affiliations:** Center for Environmental Biotechnology, University of Tennessee, Knoxville, TN, United States; Department of Microbiology, University of Tennessee, Knoxville, TN, United States; Department of Civil and Environmental Engineering, University of Tennessee, Knoxville, TN, United States; Department of Biosystems Engineering and Soil Science, University of Tennessee, Knoxville, TN, United States; Biosciences Division, Oak Ridge National Laboratory, Oak Ridge, TN, United States; Omni International Inc, A PerkinElmer Company, Kennesaw, GA, United States; Michigan State University College of Osteopathic Medicine, East Lansing, MI, United States; Jeevan BioSciences, Tucker, GA, United States; Department of Biology, Georgia State University, Atlanta, GA, United States; Battelle Memorial Institute, Columbus, OH, United States

**Keywords:** SARS-CoV-2, variant of concern, diagnostics, sample preparation

## Abstract

The SARS-CoV-2 pandemic has profoundly impacted communities across the globe, requiring accurate and accessible diagnostic technologies in support of public health mitigation efforts. As testing has evolved throughout the course of the pandemic, varying sample preparation methodologies have been employed. Herein we perform a comparison of three commercial sample preparation methods: two mechanical homogenization workflows and one enzymatic digestion approach for the detection of SARS-CoV-2 from biomarker genes in 20 human saliva pools. SARS-CoV-2 variants of concern were also identified on the University of Tennessee, Knoxville campus during the spring semester of 2021 utilizing the commercial PerkinElmer PKamp VariantDetect SARS-CoV-2 RT-PCR Assay kit. Two hundred and ten (210) human saliva pools were selected and analyzed for the presence of SARS-CoV-2 variants of concern providing insight into the utility of these various commercial workflows for integration into current public health SARS-CoV-2 surveillance measures.

## Introduction

Accurate and accessible diagnostic testing provides the cornerstone of most public health and treatment efforts for viral disease. The global pandemic caused by SARS-CoV-2 infections required rapid advancement in diagnostic techniques and technologies to maintain pace with the global demand for accurate viral testing [1]. Throughout the pandemic, quantitative polymerase chain reaction (qPCR) based diagnostic assays have been the gold standard for SARS-CoV-2 diagnostics [1]. In March 2020, ThermoFisher Scientific commercialized one of the first diagnostics assays for the detection of SARS-CoV-2; a TaqPath COVID-19 qPCR based kit which relies on fluorescence technologies allowing for multiplexing of PCR assays to detect and distinguish between different gene targets in the same sample. The TaqPath COVID-19 qPCR kit was granted emergency use for clinical testing by the FDA on March 13, 2020 (https://www.fda.gov/media/136113/download). The qPCR based assays developed for SARS-CoV-2 diagnostics have experienced multiple iterations of refinement, evolving from the initial nasopharyngeal swab tests to assays for other sample materials including saliva, wastewater, and feces [2–6]. This rapid diversification of sample inputs for qPCR detection of SARS-CoV-2 biomarkers was driven by a necessity to maintain public compliance with testing efforts while maintaining accuracy and availability of testing.

As the pandemic progressed, multiple variants of concern emerged across the globe with the potential to evade current diagnostic assays and available treatments [7]. It soon became a priority of many health organizations to identify and track these emerging variants of concern across different geographic areas and populations [8]. Diagnostic assay development became an increasing priority to allow for SARS-CoV-2 variants of concern detection to support public health efforts.

In this study, we compare three qPCR based methodologies for SARS-CoV-2 diagnostics using human saliva samples collected on the University of Tennessee, Knoxville campus. Two of these methodologies, the SalivaDirect with Proteinase K and the KingFisher RNA Extraction method, are commercially available and were compared to the Omni Direct-to-PCR (dPCR) method [9–11]. This comparative evaluation provides insight into the ability of different workflows to accurately detect viral RNA within saliva samples. While less invasive for the patient to provide, these samples often have increased PCR inhibitors present, making saliva a more difficult sample to process and achieve sensitive detection of SARS-CoV-2 biomarker genes. Thus, comparative studies assessing the impact of sample preparation workflows are crucial for establishing confidence in diagnostic testing with this sample medium.

Additionally, the human saliva samples collected for this study were screened for variants of concern utilizing the commercial PerkinElmer PKamp VariantDetect SARS-CoV-2 RT-PCR Assay kit. These samples were collected throughout the 2021 spring semester, and demonstrated the ability of this kit to accurately detect and differentiate between variants of SARS-CoV-2 as they spread throughout the campus community.

### Materials and Methods

This study assessed the efficacy of the Omni protocol for detection of SARS-CoV-2 RNA in de-identified human saliva samples. The performance of the Omni Direct-to-PCR (dPCR) approach was compared against a standard RNA extraction method (i.e., KingFisher extraction) commonly used in Clinical Laboratory Improvement Amendments (CLIA) certified labs as well as the SalivaDirect protocol developed by the Yale School of Public Health, New Haven, CT [12]. The Omni dPCR method utilizes samples homogenized by the Bead Ruptor Elite (Omni, Cat. No. 19-042E) and then added directly to the qPCR reactions. The KingFisher extraction protocol relies upon viral particle lysis and mechanical disruption of the virus. The SalivaDirect protocol combines both Proteinase K and heat treatment to denature human saliva samples.

The samples utilized in this study were obtained from the University of Tennessee, Knoxville campus in the Spring 2021 semester. All participants were consented via written and oral consent prior to providing saliva samples. Saliva samples provided for the research in this study were deidentified prior to being handed over to the lab for further use. All research was approved under protocol IBC-20-547-2 approved on July 6 2020 by the UTK institutional review board.

In early summer 2020, the University of Tennessee, Knoxville campus developed and validated protocols to detect SARS-CoV-2 for public health surveillance using RT-qPCR assays that target the viral envelope (E), nucleocapsid (N), RNA polymerase (RdRP), and human Ribonuclease P (RNase P, used as a positive control) genes for human saliva under IBC-20-547-2 (IBC-20-547-2; July 6, 2020). The E gene provides for the outermost layer of the virus protecting the genetic material when traveling from one host to another. The N gene provides the protective coat of proteins associating with the viral nucleic acids. The RdRP gene is one of 16 nonstructural proteins that are produced in the overlapping open reading frames (ORF1ab region) that encode for polyproteins PP1ab and PP1a. The function of these nonstructural proteins includes viral transcription, replication, proteolytic processing, suppression of host immunity and host gene expression [13]. The E, N, and RdRP/ORF1abregions have been the commonly analyzed viral targets for diagnosis.

The de-identified human saliva samples used in this study were chosen based upon previous testing using the protocols established by the University of Tennessee SARS-CoV-2 Surveillance Testing Laboratory to include known SARS-CoV-2 positive and known SARS-CoV-2 negative human saliva samples. Table 1 below shows the published primer and probe sets specifically targeting the E, RdRP, and RNase P genes that were used in this study [3, 4, 6]. Although the viral N gene was originally included in the multiplexed qPCR assays during previous testing, it is excluded in this study as both the E and RdRP targets are sufficient for detecting the presence of the viral genome in the saliva samples.

**Table 1.**
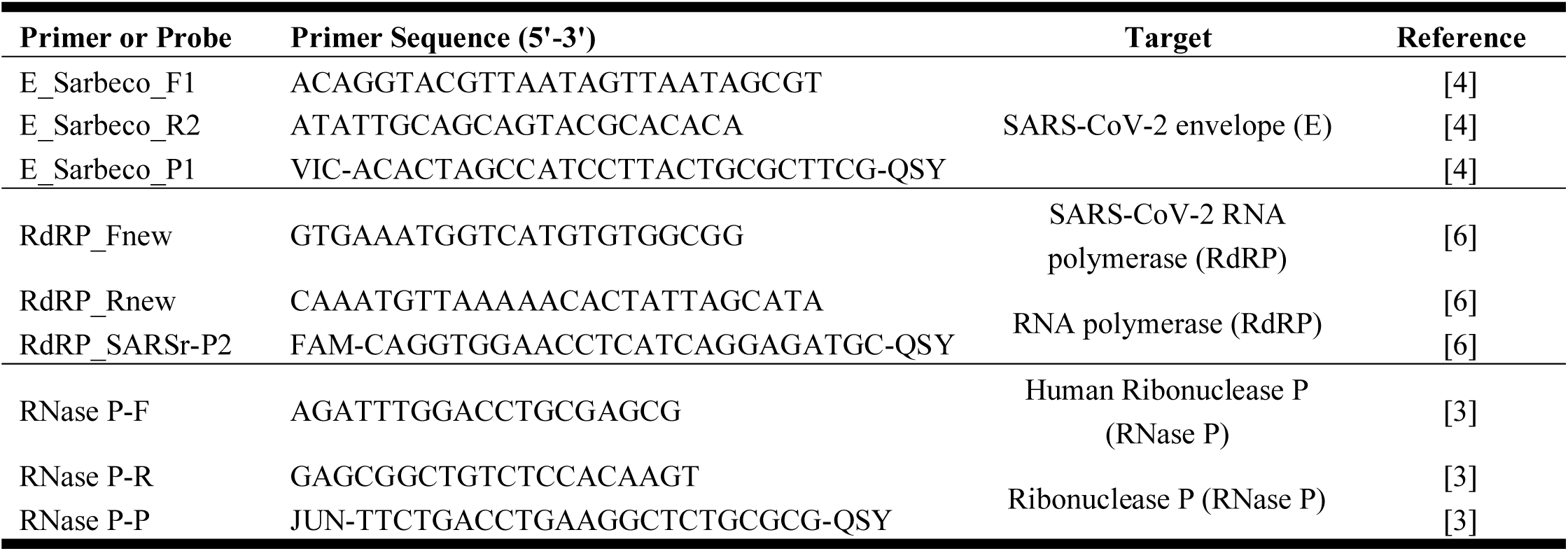
Primers and probes used in this study.

Omni provided a PerkinElmer PKamp VariantDetect SARS-CoV-2 RT-PCR Assay kit (PerkinElmer, Cat. No. 4224-0010) to detect and differentiate SARS-CoV-2 variants. The PerkinElmer kit uses different combinations of multiplexed primers and TaqMan probes to detect different SARS-CoV-2 variants (i.e., combination C detects wildtype, Alpha, Beta and Gamma variants and combination F detects Delta and Kappa variants). To demonstrate the utility of this kit, 210 de-identified human saliva pools were used to test the validity and efficacy of this kit. To verify that assay reagents performed according to expectations, the controls used included an extraction control with nuclease free water added instead of a human saliva sample; a no template PCR control with nuclease-free water added instead of RNA during RT-qPCR; and a positive control of 5 μL of 1.05×10^3^ genome copy equivalents/μL of RNA from inactivated SARS-CoV-2 (USA-WA1/2020) was added during RT-qPCR (BEI Resources, Cat. No. NR-52286). The PKamp VariantDetect SARS-CoV-2 RT-PCR Assay kit detects 4 different SARS-CoV-2 mutations which include the Alpha variant (B1.1.7 – United Kingdom, September 2020), the Beta variant (B.1.351 – South Africa, May 2020), Gama variant (P.1 – Brazil, November 2020), Delta variant (B.1.617 – India, October 2020), and the Kappa variant (B.1.617.1 – India, April 2021) (Table 3).

### Experiment 1: Comparison of Sample Processing Method

Twenty saliva pools each consisting of five individual human saliva samples were selected including 18 known positive (+) saliva pools; and 2 known negative (-) saliva pools. TaqPath™ 1-Step Multiplex Master Mix (No ROX) (ThermoFisher Cat. No. A28523; Waltham, MA) was added to multiplex assays (Table 2) for the detection of the E gene, the RdRP gene, and the RNase P gene, the latter serving as an internal amplification control gene. Extraction negative controls were established by adding sterile water in the place of a saliva pool. RT-qPCR negative controls (no template control) consisted of complete assays with 5 μL of nuclease-free water replacing the RNA template. Positive controls for the E and RdRP genes were established by adding 5 μL of 1.05×10^3^ genome equivalents/μL synthetic RNA to give a concentration of 5.25×10^3^ genome copy equivalents per reaction (BEI Resources, Cat. No. NR-52286).

**Table 2.**
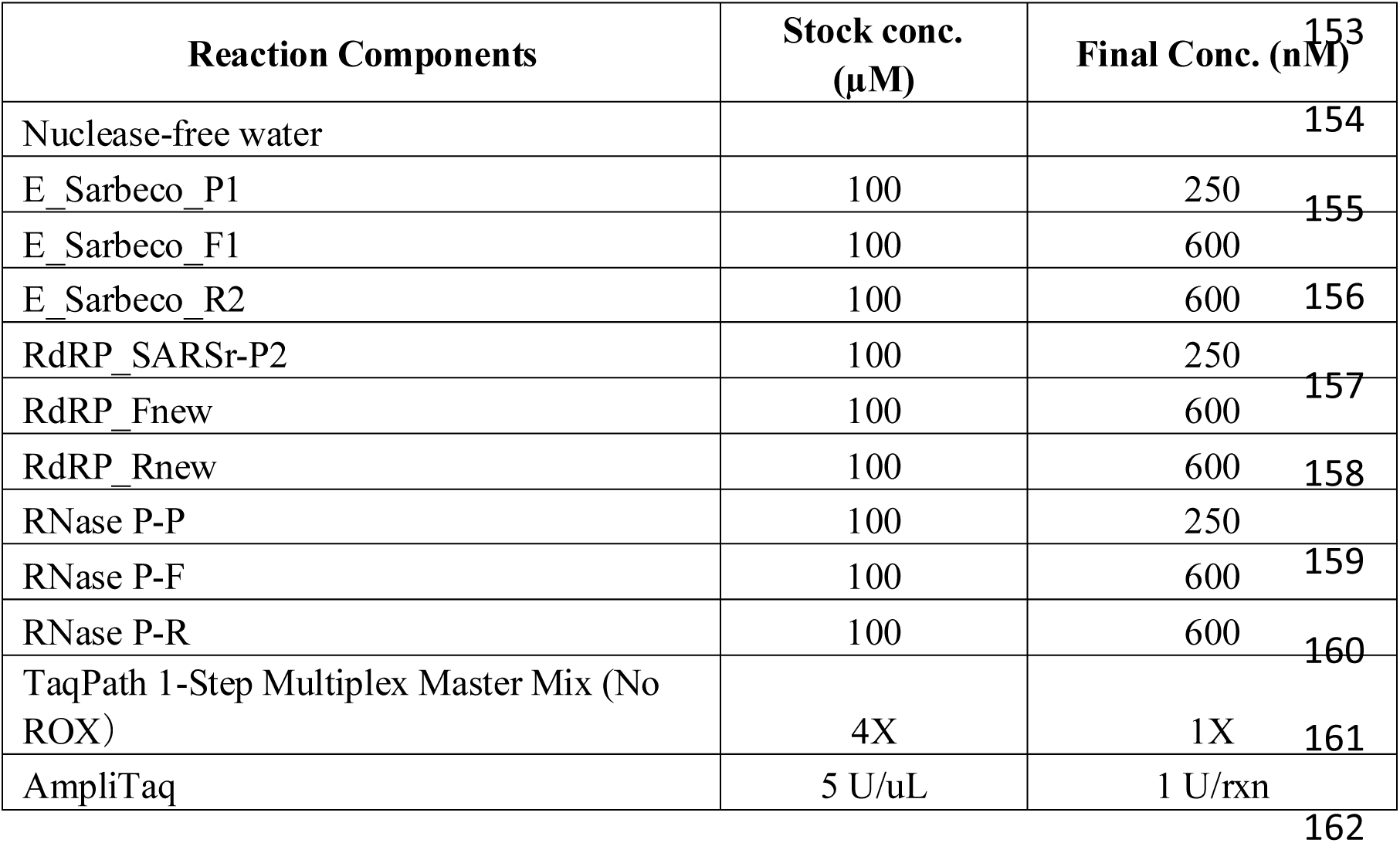
Rt-qPCR reaction multiplex assay mixture used for each of the three processing methods with the only change being the source of the template.

1. <u>Omni Direct-to-PCR method:</u> Each human saliva pool (500 μL) was transferred to a 2-mL Omni bead tube (Omni, Cat. No. 19-628). The bead tubes were placed inside the Bead Ruptor Elite (Omni, Cat. No. 19-042E) and agitated at 4.5 m/s for 30 seconds. Five μL of each processed pool were used for qPCR.
2. <u>KingFisher extraction method:</u> The MagMAX™-96 Viral RNA Isolation Kit and kit supplied buffers were used for this extraction method following manufacturer protocol (ThermoFisher, Cat. No. AM1836; Waltham, MA) with the addition of Dithiothreitol (DTT) to lower the viscosity of the saliva. DTT (Sigma-Aldrich, Cat. No. 501656980, Inc., St. Louis, MO) (20 μL of a 50 mM aqueous stock solution) was added to 200 μL of each human saliva pool and incubated at room temperature for 30 minutes. After the incubation, Proteinase K (5 μL of a 1-5% aqueous stock solution; ThermoFisher Cat. No. A42363; Waltham, MA) and 275 μL binding buffer/bead mix (provided in kit) were added to each sample. The RNA was first washed with wash buffer (provided in kit) and then with 80% molecular grade ethanol, before RNA was eluted in 50 μL elution buffer (provided by in kit). Five μL of the RNA extract from each sample were used for qPCR
3. <u>RNA extraction following SalivaDirect with Proteinase K treatment:</u> Proteinase K (2.5 μL of a 1-5% aqueous stock solution; ThermoFisher Cat. No. A42363; Waltham, MA) was added to 50 μL of human saliva pool and vortexed for 1 minute. Samples were then heated to 95°C in the Veriti™ 96-well Thermal Cycler (Applied Biosystems, Model No. 9902, Waltham, MA) for 5 minutes. Five μL from each sample were used for qPCR [12].

### Experiment 2: Variant tracking

A total of 210 positive human saliva pools collected from September 2020 through April 2021 were subjected to variant testing with the Pkamp VariantDetect SARS-CoV-2 RT-PCR Assay kit. These 210 positive human saliva pools were selected based upon previous identification using the multiplex qPCR assay protocol established by the University of Tennessee SARS-CoV-2 Surveillance Testing Laboratory. RNA was freshly extracted from each of the 210 positive human saliva pools using the KingFisher Flex MagMAX™-96 Viral RNA Isolation Kit per manufacturer protocol (ThermoFisher, Cat No. AM1836; Waltham, MA) with the addition of DTT. Immediately following RNA extraction, RT-qPCR was performed using the PKamp VariantDetect SARS-CoV-2 RT-PCR Assay kit and the corresponding protocols provided by the manufacturer. The PKamp VariantDetect SARS-CoV-2 RT-PCR Assay kit combination C (which detects wildtype, Alpha, Beta and Gamma variants) was used to identify variants present in these 210 pools. If combination D testing did not clearly identify the variant present in a pool, that pool was then tested using combination F (which detects Delta and Kappa variants).

## Results

Experiment 1 compared the three different human saliva pool processing methods, and the results are shown in Figure 1. The RNase P gene was amplified from all pools using all three processing methods. False positive results were obtained for the E gene in negative pools when the Omni Direct-to-PCR method was used (Figure 1 and Figure 2). These samples were determined to be false positives as they were initially tested following the established protocols by the University of Tennessee SARS-CoV-2 Surveillance Testing Laboratory and were known to be negative for SARS-CoV-2. Of note is the observation that, in a small proportion of the 210 human saliva pools, the RdRP gene was not amplified with the assay that targets the ORF1ab region (Figure 1). Amplification of the RdRP gene was only observed in pools processed when the KingFisher extraction method was used.

**Figure 1.**
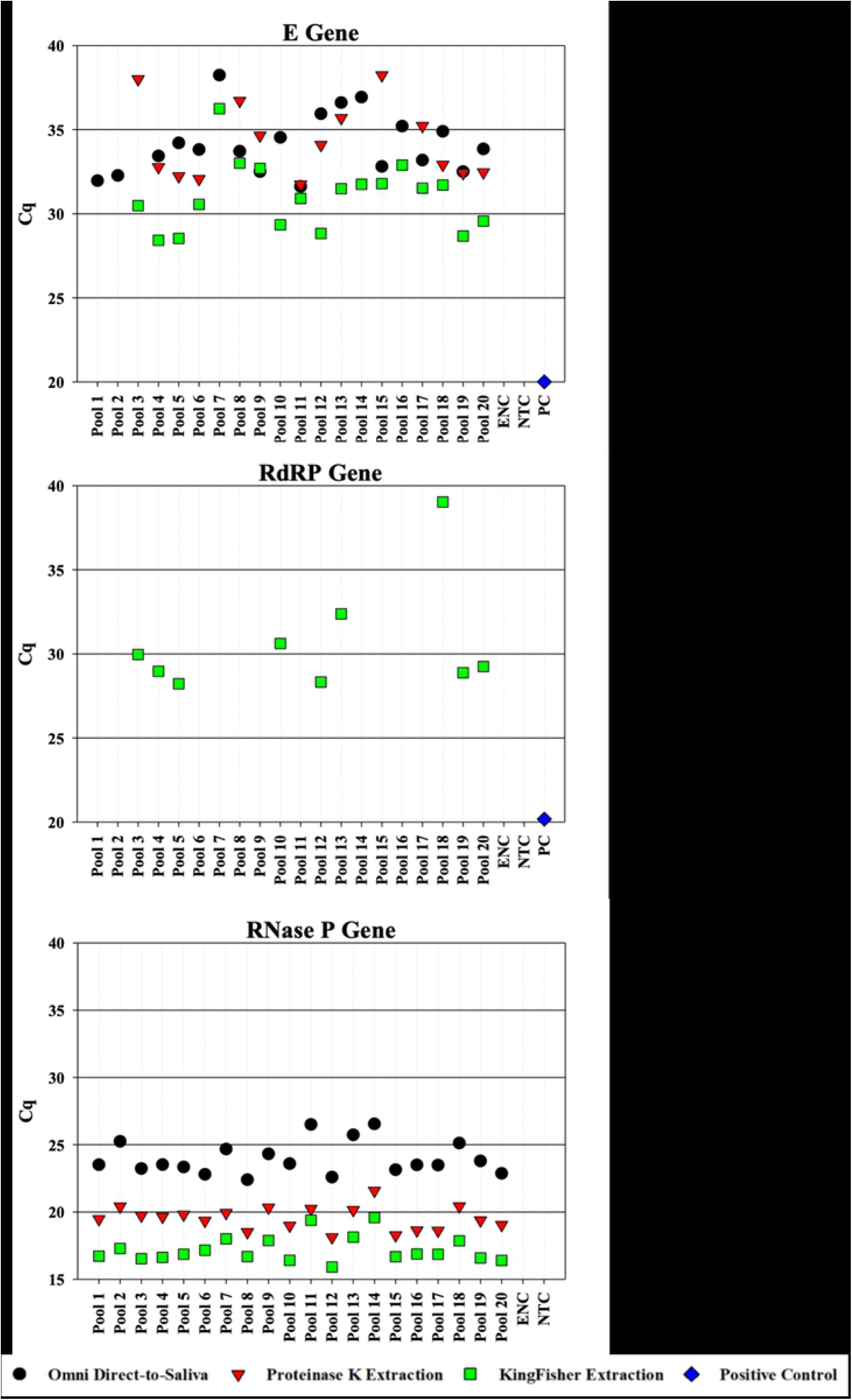
qPCR assays targeting the E, RdRP and RNase P genes in 20 sample pools plus controls. No amplification occurred in assays targeting the RdRP gene of the ORF1ab region of the RdRP gene of SARS-CoV-2. The negative saliva pools are Pool 1 and Pool 2. Extraction Negative Control (ENC); No Template Control (NTC); Synthetic SARS-CoV-2 RNA (PC).

**Figure 2.**
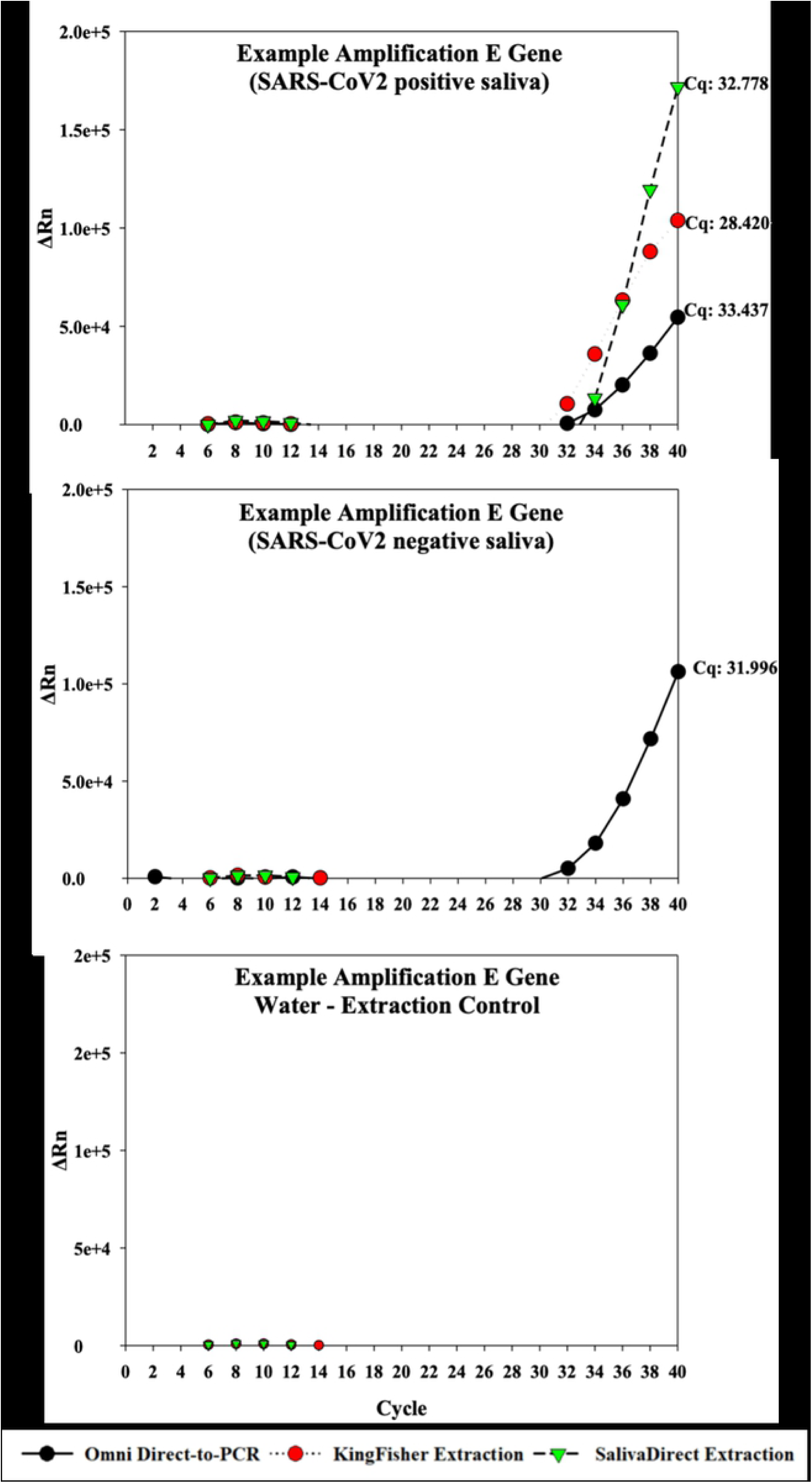
Representative linear qPCR amplification plots for the E Gene showing false positive amplification seen in SARS-CoV-2-negative samples with RNA extracted using the Omni Direct-to-PCR method. The Omni Direct-to-PCR method using known negative saliva amplifies with a Cq of 31.966, which is earlier than the known positive saliva with a Cq of 33.437. The Positive sample example is Pool 4 and the negative sample is Pool 1.

Figure 2 illustrates representative linear RT-qPCR amplification plots showing the SARS-CoV-2 negative human saliva pool with the E gene amplifying earlier than in the SARS-CoV-2 positive human saliva pool using the Omni RNA extraction method. Figure 2 demonstrates the false positive amplification in the known SARS-CoV-2 negative human saliva using the Omni dPCR method. Both the known SARS-CoV-2 positive human saliva pool and the known SARS-CoV-2 negative human saliva pool using the Omni dPCR method resulted in Cq value above 30. The SalivaDirect extraction method and the Omni dPCR method both amplified higher than a Cq value of 30 but the SalivaDirect extraction method did not show amplification in the known SARS-CoV-2 negative human saliva pool. Four independent replicate experiments using different reagents each time showed the same results.

Experiment 2 tracked the variant distribution of SARS-CoV-2 that occurred during the spring semester of 2021 at the University of Tennessee, Knoxville campus. Table 3 outlines the various deletions/mutations found in the variants of interest detected by PKamp VariantDirect SARS-CoV-2 RT-PCR Assay kit with Table 4 showing the results of the variant tests for the pools used in this study. Figure 3 demonstrates how the SARS-CoV-2 variant prevalence changed over time. Early in the spring semester (mid-March), almost all positive pools were caused by the original wild type of the SARS-CoV-2 virus. Mid to late spring semester (mid-March to the end of April), a shift in the SARS-CoV-2 variant was observed from wild type to mostly the Alpha variant (UK strain). Of the 210 human saliva pools subjected to variant testing, only 18 pools failed to be identified with the PKamp VariantDetect SARS-CoV-2 RT-PCR Assay kit combinations used in this study. The mutations of the SARS-CoV-2 variants found in the 18 pools that failed to be identified with the PKamp VariantDetect SARS-CoV-2 RT-PCR Assay kit combinations are listed in Table 4.

**Table 3.**
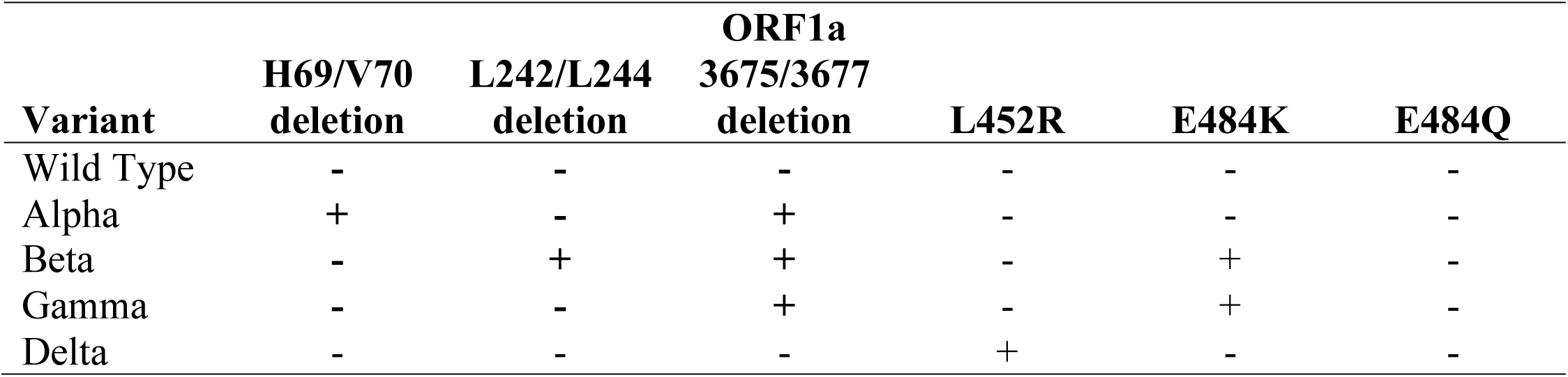
SARS-Cov-2 variants showing the deletions characteristic for each variant [reference? where did this information come from?]. Presence of the mutation is indicated by (+) and absence of the mutation is indicated by (-).

**Table 4.**
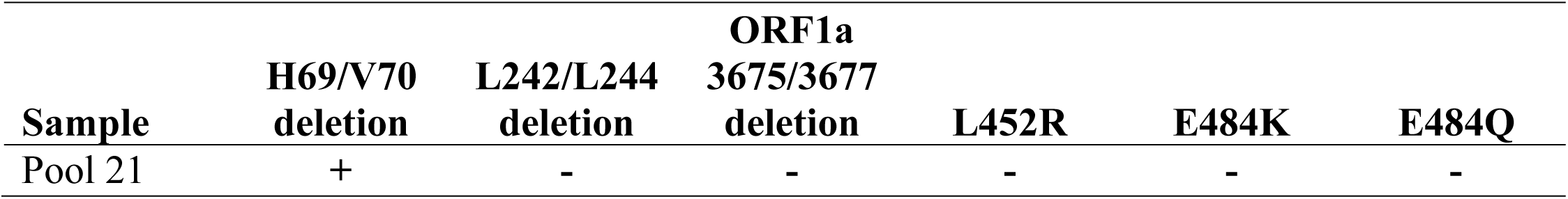
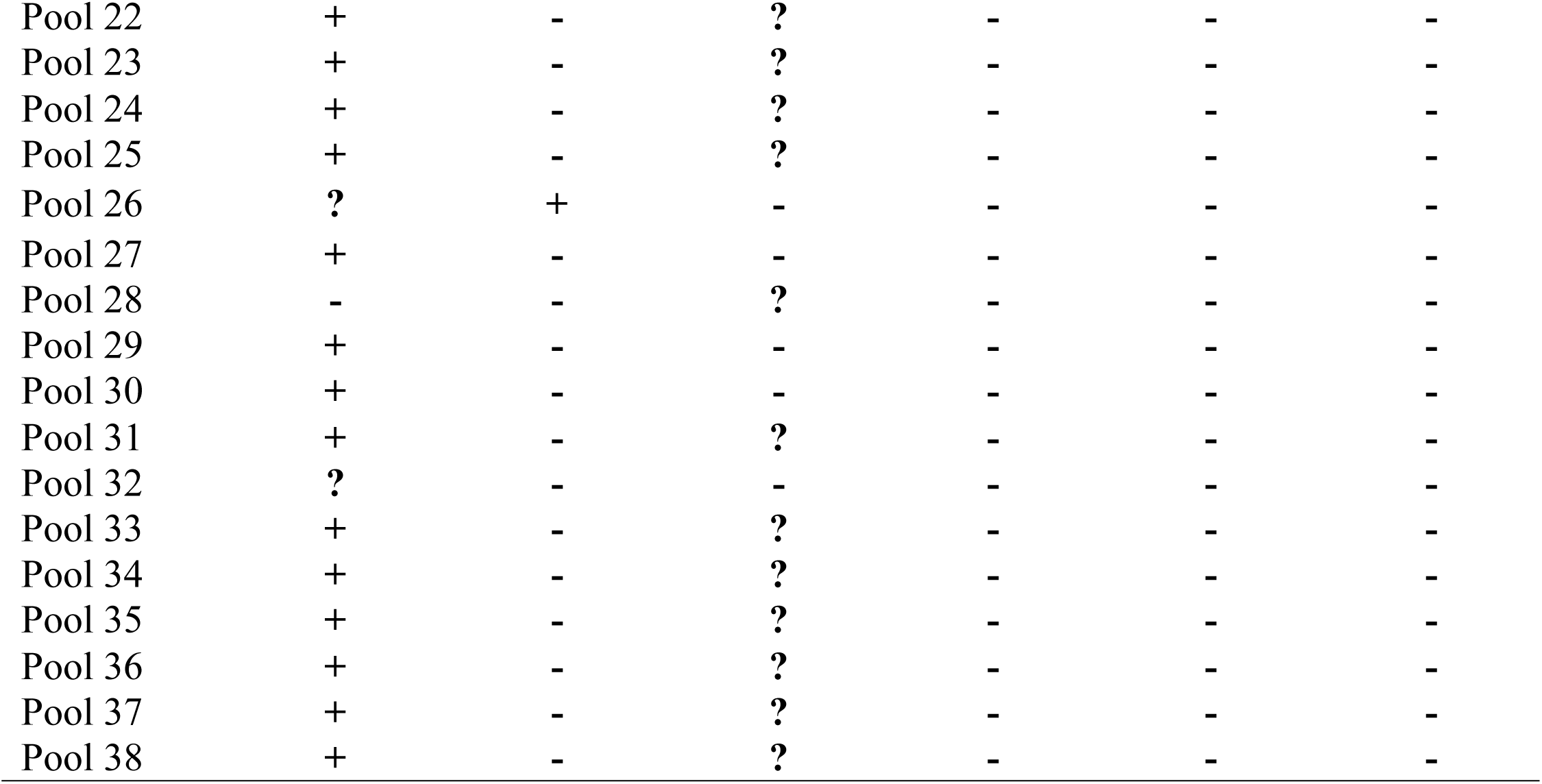
Mutations present in samples with unidentified variants after both combinations D and F of the PKamp VariantDetect SARS-CoV-2 RT-PCR assay kit tests were performed. Presence of the mutation is indicated by (+) and absence of the mutation is indicated.

**Figure 3.**
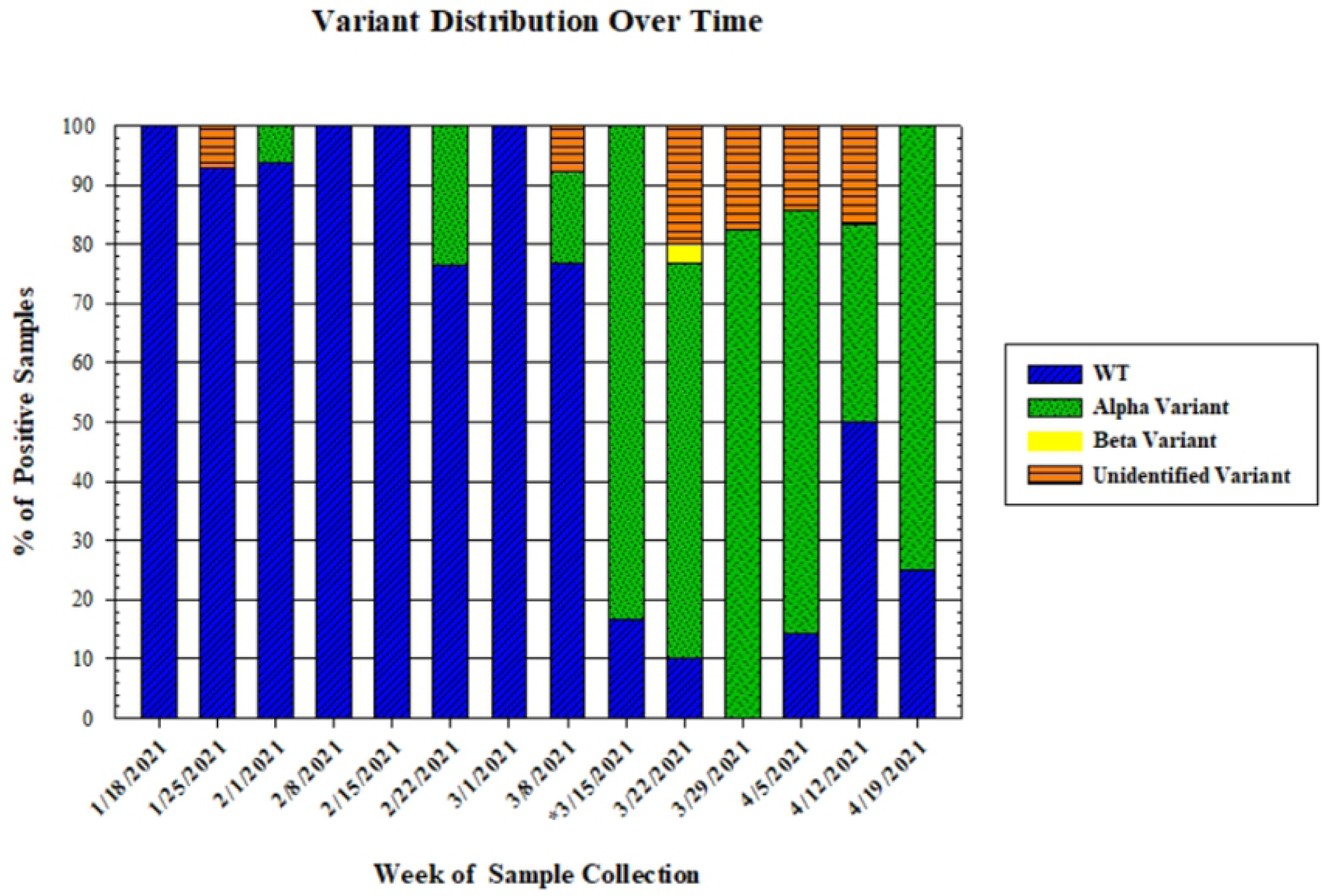
SARS-CoV-2 variant distribution across the University of Tennessee campus in spring 2021. The asterisk (*) depicts the week that spring break usually occurs.

## Discussion

As illustrated in Figure 2, we noted that the Omni Direct-to-PCR (dPCR) method resulted in false positive amplification of the SARS-CoV-2 E gene in known SARS-CoV-2 negative human saliva pools. Both the known SARS-CoV-2 positive pool and the known negative pool using the Omni dPCR method showed amplification while the SalivaDirect extraction method did not show amplification in the known negative human saliva pool. A plausible explanation of the observations is nonspecific amplification or artifacts in the known SARS-CoV-2 negative human saliva that cause the observed false positive results. These artifacts can be caused by an imbalance between primers, template and non-template RNA concentrations [14]. The negative control for the Omni dPCR method showed no amplification, which leads us to attribute the amplification of the E gene in the SARS-CoV-2 negative human saliva samples to the Omni dPCR method itself. The test was repeated 4 separate times with new reagents each time to assure contamination was not the issue. Each repetition produced similar results. Contamination of the kit was ruled out as a contributing factor from the manufacturer due to the quality control metrics reported wherein, they demonstrated the absence of any RNA in the kits via PCR and gel visualization prior to shipment.

In Experiment 2, the PKamp VariantDetect SARS-CoV-2 RT-PCR Assay kit identified which variant was present in the majority of human saliva pools collected on the University of Tennessee, Knoxville campus during the spring of 2021. These results demonstrate the ability of the variant detection kit to quickly detect and differentiate between SARS-CoV-2 variants.

Although we cannot make definite conclusions on the variants present in these 18 SARS-CoV-2 positive human saliva pools listed in Table 4, we are able to gain some valuable information. The majority of these 18 pools show H69/V70 deletions which is characteristic of the Alpha variant lineage. The ORF1a 3675/3677 deletion, which was not detected or did not meet kit specifications, may be in a region of the gene that has been speculated to have higher mutation rates. Mutations found in the S-protein and the ORF1a and ORF1b genes can alter the viral attachment, fusogenicity and/or immunogenicity as well as interrupt the viral proofreading [15, 16]. It has been suggested that some mutations in the ORF1a and ORF1b regions that alter the viral replication can increase down-stream mutations [15, 16]. By piecing this information together, we can hypothesize that there may be a SARS-CoV-2 variant of the Alpha lineage, with increased rates of mutation present in our sampling community. By monitoring the rate of these mutations in the population, we can monitor the rate of transmission of this particular lineage and potentially detect upcoming variants of interest.

It is interesting to note that there were no confirmed cases of the Delta variant on the University of Tennessee, Knoxville campus during the testing period. Considering the increased transmissibility, this variant was expected to be prevalent in the later months of the spring semester. Future studies could investigate the timeline of this variant in surrounding communities and allow a better understanding of the spread of the Delta variant on campus.

Before the PKamp VariantDetect SARS-CoV-2 RT-PCR Assay kit combo F kit was available, we attempted to identify variants using the GT Molecular RT-qPCR SARS-CoV-2 Delta Variant Mutational Signature Assay Kit (Fort Collins, CO; SKU: 100172). We did not observe amplification using this kit with any of the human saliva samples collected on campus.

Herein we successfully evaluated three qPCR-based assays for the detection of SARS-CoV-2 from saliva pooled samples obtained across the University of Tennessee, Knoxville campus in early 2021. While evaluating the three methods, the Omni Direct-to-PCR method, SalivaDirect, and the King Fisher extraction method, they were shown to have a greater than 80% agreement between all methods when identifying SARS-CoV-2 positive saliva pools. Secondary to the successful identification of SARS-CoV-2 in the pooled saliva samples, it was shown that the mechanical homogenization technique utilized by the Omni dPCR method provided sufficient viral lysis and exposure of the viral RNA to go directly into qPCR detection, bypassing traditional extraction methods while maintaining sensitivity in comparison to two other extraction-based methods evaluated. In addition to the evaluation of the three qPCR assays, the saliva pools were tested using the PKamp VariantDetect SARS-CoV-2 RT-PCR Assay to track the presence and proportionality of multiple variants of concern across the university’s campus during the spring 2021 semester. When evaluating variants, an observed shift from wild type to alpha variant was seen surrounding spring break. This shift corresponds with a national trend seen at that time as the alpha variant transitioned to becoming the dominant variant across the US. An unidentified variant was seen as the semester progressed, but it is likely that that is the emergence of a recombination seen following spring break with large portions of the student body traveling and was an event that did not correspond with a variant of concern identified in the utilized assay. Further evaluation of the unidentified variant could be considered for future studies on the genomic diversity of SARS-CoV-2 across university populations. Overall, this manuscript successfully demonstrates the ability of multiple commercially available assays to successfully utilize pooled saliva samples for highly sensitive SARS-CoV-2 detection.

## Data Availability

All data pertaining to this study is presented and available in the manuscript without alterations or restrictions.

## Acknowledgments

We acknowledge the University of Tennessee Office of Research and Engagement for providing funding for this project and Omni International Inc, A PerkinElmer Company. Additionally, we would like to acknowledge and thank Mr. Pete Tortorelli of Omni International for his continued leadership and financial support of our research endeavors.

## Author Contributions

Author contributions to this manuscript have been determined utilizing the CRediT criteria as denoted below. **MR Poindexter:** conceptualization, data curation, formal analysis, investigation, visualization, writing – original draft, writing – review and editing. **T Xu:** data curation, formal analysis, methodology, investigation, visualization, writing – original draft, writing – review and editing. **CM Swift:** data curation, formal analysis, visualization, writing – original draft, writing – review and editing. **CM Proctor:** investigation, methodology, writing – review and editing. **F Kara-Murdoch:** formal analysis, validation, writing – review and editing. **ZP Morehouse:** conceptualization, methodology, writing – original draft, writing – review and editing. **GL Ryan:** methodology, writing – review and editing. **FE Löffler:** conceptualization, formal analysis, funding acquisition, investigation, methodology, project administration, resources, supervision, writing – review and editing. **RJ Nash:** conceptualization, funding acquisition, investigation, methodology, project administration, resources, supervision, writing – review and editing.

